# Controlled Avalanche – A Regulated Voluntary Exposure Approach for Addressing Covid-19

**DOI:** 10.1101/2020.04.12.20062687

**Authors:** Eyal Klement, Alon Klement, David Chinitz, Alon Harel, Eyal Fattal, Ziv Klausner

**Affiliations:** Koret School of Veterinary Medicine, Department of Epidemiology and Public Health, the Hebrew University, Jerusalem, Israel; Buchmann Faculty of Law, Tel-Aviv University, Israel; Department of Health Policy and Management, School of Public Health, Hebrew University, Jerusalem, Israel; Phillip and Estelle Mizock Chair in Administrative and Criminal Law, The Faculty of Law, the Hebrew University, Jerusalem, Israel; Department of Applied Mathematics, Israel Institute for Biological Research, Ness-Ziona, Israel

## Abstract

**Background:** The ongoing Covid-19 pandemic has driven many countries to take radical suppression measures. While reducing mortality, these measures result in severe economic repercussions, and inhibit the development of herd immunity. Until an effective vaccine will be available, we propose an alternative approach, akin to avalanche control at ski resorts, a practice which intentionally triggers small avalanches in order to prevent a singular catastrophic one. Its main goal is to approach herd immunity faster than the current alternatives, with lower mortality rates and lower demand for critical health-care resources. According to this approach, individuals whose probability of developing serious health conditions is low (i.e. 20-49 years old with no comorbidities) will be offered the option to be voluntarily exposed to the virus under controlled supervision, and will then be issued ‘immunity certificates’ if they are confirmed to have developed SARS-CoV-2 antibodies.

**Methods:** Using a compartmental model we examine the implications of the controlled avalanche (CA) strategy over the population in Israel. We compare four scenarios: in two scenarios the CA program is applied to the low-risk population (with the rest of the population subject to mitigation measures), followed by mitigation for the entire population or by uncontrolled spread. These are compared to mitigated and uncontrolled scenarios without the CA program. We discuss the economic, ethical and public health implications of the CA strategy.

**Findings:** We show that compared to mitigation of the entire population, the CA strategy reduces the overall mortality by 43%, reduces the maximum number of people in need for ICUs by 62% and decreases the time required for release of 50% of the low-risk population by more than 2 months.

**Interpretation:** This study suggests an ethically acceptable practice, that enables reaching herd immunity faster than the current alternatives, with low mortality and minimal economic damage.

## Introduction

The ongoing Covid-19 pandemic has created a crisis where optimal control and means of mitigation are still unknown. Two main competing strategies have been suggested and implemented for controlling the epidemic: The first, *suppression*, aims at drastic reduction of disease incidence. This strategy is based on radical social distancing, closure of schools and universities and forbidding mass gatherings. Policies reflecting this strategy were first enacted by the Chinese government and, subsequently, by other countries. The second strategy, *mitigation*, is based on case isolation, voluntary home quarantine, and social distancing of high-risk populations. This strategy was successfully implemented in Israel during the first three weeks of the epidemic. However, it was abandoned and replaced by a suppression strategy due to a sharp increase of disease incidence following a holiday **(1)**. Mitigation strategy was also practiced for a short time in England before being abandoned after Ferguson et al. demonstrated their projections **(2)** and it is currently practiced in Sweden.

Ferguson et al., **(2)** demonstrated that although mitigation would reduce peak critical health care demand, it would still be several times higher than its surge capacity. Consequently, it would result in a staggering death toll. These grim forecasts were realized in the epidemics in Italy, Spain, France and the US, which shifted to suppression only during late stages of the epidemic. Suppression, on the other hand, though effective in controlling the epidemic, would prevent the development of herd immunity and is expected to paralyze the economy and carries far-reaching social ramifications.

Some countries (e.g. Taiwan, South-Korea and Singapore) have followed WHO recommendations (3), and implemented social distancing with contact tracing accompanied by massive testing and isolation of the infected prior to symptoms. These countries have been relatively successful in mitigating the epidemic and reducing case fatality so far (4), though none of them has eliminated the virus.

Covid-19 is characterized by a high R_0_, probable infectiousness of asymptomatic and pre-clinical patients (5), and has no known cure, precluding containment (6). Therefore, in the absence of effective vaccine, the only foreseeable endpoint of this epidemic can be expected when a certain level of herd immunity is reached. Such a condition requires a large proportion of the population (58%-70%) to be infected, depending on the estimated R_0_ (2.4-3.3) (2, 7). The main goal of any control strategy should be therefore to reduce the percentage of high-risk population among those infected until herd immunity is reached, and use critical health care resources optimally over time.

The strategy we propose is based on identifiable risk characteristics of different segments in the population and on voluntary immunization through exposure. Instead of using non-discriminating measures targeted at the population as a whole, we propose regulated voluntary exposure of its low-risk members. Once they are certified as immune, these individuals return to the population, increase its overall immunity and resume their normal life. This approach is akin to avalanche control at ski resorts, a practice which intentionally triggers small avalanches in order to prevent a singular catastrophic one. Its main goal is to create herd immunity, faster than current alternatives, and with lower mortality rates and lower demand for critical health-care resources. Furthermore, it is also expected to be effective in relieving the huge economic pressures created by the current pandemic (For similar proposals see supplementary material).

Using a compartmental model of the SEIRD framework we examine three main effects of the proposed strategy: total mortality, impact on the ICU at the peak of the epidemic and time of return to work. We show that all three variables are significantly reduced by the proposed strategy. We then discuss the economic, ethical and distributional aspects of its implementation. We conclude that in the current situation, this approach is reasonable on all grounds and that its underlying premises should be further examined to allow its implementation.

## Materials and methods

### 1. The Proposed Approach: Regulated Voluntary Exposure Based on Risk Characteristics

#### A. Underlying modelling assumptions

The proposed strategy dynamics was modeled, based on three assumptions:

The first assumption is that people infected by SARS-CoV-2 become immunized to future re-infection for a long time period. Previous studies demonstrated a sustained neutralizing antibody activity, lasting for at least two years, after the onset of symptoms in most of SARS-Cov-1 recovered patients (8, 9) and persistence of anti-SARS-Cov-1 IgG in infected health-care workers for at least 12 years (10). In a recent study researchers failed to re-infect Macaque monkeys 28 days after first clinical infection, indicating protective immunity developed after infection by SARS-Cov-2(11).

The second assumption, based on experience with SARS-Cov-2 is that the period of infectiousness is limited and that virus shedding can be easily detected. Data collected so far suggest that virus shedding by moderately affected Covid-19 patients does not exceed 14 days after onset of clinical signs (12, 13). Virus shedding was documented in asymptomatic patients for at least seven days (14). Though there is no documentation of the longest period of virus shedding in asymptomatic patients it is very reasonable to conclude that they do not shed the virus longer than cases who show clinical signs. In accordance with this evidence, the current guidelines of the ECDC and other health agencies for hospital discharge are based on resolution of clinical signs (i.e. no fever for at least 3 days and significant improvement of respiratory symptoms combined with at least two negative consecutive respiratory tract samples taken at least 24 hours apart (14)).

The third assumption is that case fatality among people aged 20-_49_ years is very low. According to Verity et al. the proportions of cases hospitalized among patients aged 20-29, 30-39 and 40-49 in China were 1%, 3.4% and 4.3%, respectively. Case fatality ratios (CFR) estimated for these ages in Wuhan were 0.03%, 0.08% and 0.16% respectively (15). Similar age specific CFRs were reported in South-Korea. Though Verity et al. did account for age differential under-ascertainment and the Korean data is based on very meticulous surveillance, the real extent of under-ascertainment may be lower and can be estimated only after availability serologic data. It should also be considered that these case fatalities include patients with co-morbidities even in the younger ages.

#### B. Regulated voluntary exposure program

Given the above three assumptions, the following program is suggested: Individuals whose probability of developing serious health conditions is low will be offered the option to be exposed to the virus under controlled supervision. To do so, a threshold risk factor should be determined, based on easily identifiable and verifiable characteristics, most significantly age, but also the existence of other comorbidities such as hypertension, cardiac disease, lung disease, diabetes, etc.. Individuals whose risk is lower than this threshold would be allowed to participate in the voluntary exposure program.

Exposure may be applied using one of various alternative measures – either through individual controlled infection, or (as modelled here) through coordinated group exposure to the virus and quarantine, where all participants can communicate and contract the virus, thus raising the effective reproductive number within this group. It may also be implemented by simply relaxing social distancing and other limitations for the participants, subject to subsequent quarantine and periodical medical monitoring.

The state would issue ‘immunity certificates’ to everyone who is confirmed to have developed SARS-CoV-2 antibodies and is verified as non-contagious (16) (development and validation of good serological assays to detect exposure to the virus is currently defined as a major need (4) and its wide implementation is a matter of short time). Certified individuals would be allowed to reenter society and the workforce, as they can no-longer infect other people. They may also provide medical treatment, as well as other necessary services to high-risk population. These certified individuals would resume their social and economic life, and contribute to the recovery of the economy on both supply and demand sides.

### 2. Modeling the epidemiological impact

The dynamics of the COVID-19 epidemic spread as well as the effect of the controlled avalanche (CA) program were ascertained using a deterministic compartmental model which is an expansion of an earlier model (1). Similar to other models that describe the dynamics of COVID-19 in a given population, it describes the stages of the epidemic in a SEIRD framework (17-19). In this framework, the modeled population is divided into five compartments: S – susceptible, E – exposed and asymptomatic, I – infectious and symptomatic, R – recovered and immune, and D – deceased. As there is evidence for the possibility of infection by asymptomatic COVID-19 cases (5) the model consists the possibility that a fraction *ε*_*E*_ of the individuals in the E compartment can be infectious.

Due to the fact that the risk of mortality due to COVID-19 is associated with age (15), the model divides the population into two sub-populations: the low-risk labor age population that consists of individuals whose age is in the range of 20-49 and the general population which consists of all other individuals (top and middle SEIRD chains in Fig. 1, correspondingly).

**Fig. 1.**
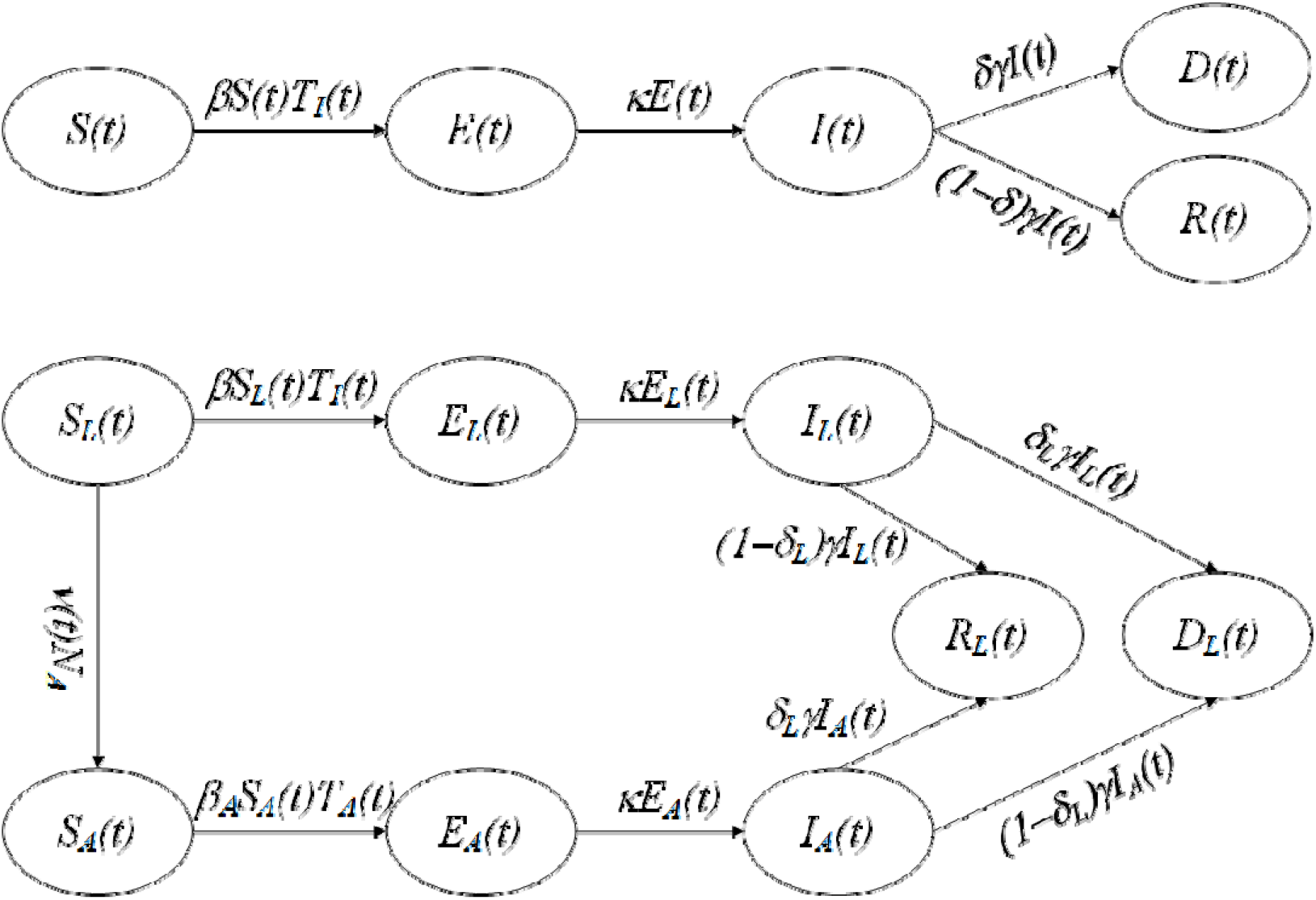
Schematic flow diagram for the Avalanche model. The model describes the Covid-19 dynamics in a given population, which is divided to five compartments: S – Susceptible, E – exposed, I – Infectious, R – Recovered and immune, D – Deceased. The population is divided to two: The general population (ages 0-19 and 50-) and the low-risk labor-age population (ages 20-49), that is indicated by L subscript. From the L population volunteers are recruited to the controlled avalanche program that is indicated by A subscript.

The low-risk and general population lead to separate dynamics. However, these two sub-populations are connected by the same group of infectious individuals, T_I_, which consists of all members of the infectious and symptomatic compartments in the general and low-risk population (namely, I and I_L_ in Fig. 1) as well as a fraction *ε*_*E*_ of the asymptomatic members of the two populations (namely, E and E_L_ in Fig. 1). Therefore, the susceptible individuals in each of the sub-populations (namely, S and S_L_) can be infected by any infectious individual from either sub-population. The disease transmission (i.e., the passage from the S to E and S_L_ to E_L_) depends also on the effective contact rate, *β*.

Following the infection, the passage of asymptomatic cases to symptomatic (i.e., from the E to I and from the E_L_ to I_L_ compartments) is dictated by the rate of development of clinical symptoms. From there, symptomatic and infectious individuals can either recover or die from the disease, according to the CFR of the general population or that of the low-risk population, *δ* and *δ*_L_, correspondingly. The passage between the I to the D or R (or from I_L_ to D_L_ and R_L_) compartments is dictated by the recovery rate, *γ*.

In the model the CA program is implemented by allowing volunteers to stay with other infected individuals at the same residency (e.g. hotels designated for the CA program), in which no social distancing is implemented and high contact rate is allowed. When the CA program is activated (low SEI chain in Fig. 1), a batch process begins. The function *υ*(*t*) is a forcing function that every *τ* days returns a value of 1, thus allowing a new batch (whose size is N_A_) of low-risk susceptible volunteers to join the CA program. In the CA program, the infection of susceptible volunteers, S_A_, is dictated by T_A_, the group of infectious individuals among the CA program volunteers and *β*_*A*_, the effective contact rate in the program. Upon recovery or death, the volunteers return to the R_L_ and D_L_ compartments of their original population.

In this study, the parameters that describe the epidemiology of the COVID-19 disease, namely *k* and *γ*, were based on an analysis of actual event-data (20). The infection fatality ratios, *δ* and *δ*_*L*_, were based on weighted average of data on Israel’s population by age groups with age dependent COVID-19 CFRs (15, 21). The periodic batch size of the CA program volunteers was determined by associated data on the inventory of active tourist hotel rooms in Israel (22). The effective contact rates, *β* and *β*_*A*_ were set according to parameters estimated for data from the COVID-19 outbreak in China, before and during control measures were taken (23, 24). Further details on the model and its parameters are provided in the supplementary part.

The dynamics of epidemic spread was modeled considering two distinct populations: low-risk (ages of 20 to 49) and the rest of the population, termed here the general population (ages 0-19 and 50-). The following four scenarios were simulated:

1. Uncontrolled epidemic spread for the entire population.
2. Mitigated epidemic spread for the entire population
3. CA program for the low-risk population with mitigation for the general population, followed by mitigation for the entire population.
4. CA program for the low-risk population with mitigation for the general population, followed by followed by uncontrolled spread.

## Results

In the modeled AC scenarios, the program was active during 210 days. During this time, the simulation included an AC participation ratio of 81.5% from the low-risk population. As depicted in Table 1, implementation of the CA policy leads to a dramatic reduction in mortality, which is accompanied by a significantly faster release of the labor population. In all scenarios, the main source of mortality is the general population. Fig. 2 demonstrates that the CA policy is fastest in releasing the lower risk - labor population, and it results in the lowest infection rate among the higher risk - general population.

**TABLE I.** Statistical parameters of the examined scenarios, regarding the general population and the low-risk labor-age population. The scenarios are: 1 -Without lockdown, 2 – effective lockdown, 3 – Controlled avalanche (CA) program during lockdown, 4 - CA program during an ongoing lockdown. At the program end, the lockdown is lifted.

| Parameter | Description | Units | Scenario 1 | Scenario 2 | Scenario 3 | Scenario 4 |
| --- | --- | --- | --- | --- | --- | --- |
| Number of deceased | General population | People | 64,299 | 48,426 | 26,405 | 47,566 |
|  | Low-risk Population | People | 2,666 | 2,008 | 2,545 | 2,538 |
|  | Total population | People | 66,965 | 50,434 | 28,950 | 50,104 |
| Maximal number of patients requiring critical care | General population | People | 35,382 | 11,839 | 4,146 | 4,146 |
|  | Low-risk population | People | 1,490 | 498 | 570 | 570 |
|  | Total population | People | 36,872 | 12,337 | 4,716 | 4,716 |
| Time of recovery of a given fraction of the low-risk population | Quarter | Days | 79 | 181 | 87 | 87 |
|  | Half | Days | 94 | 226 | 142 | 142 |
|  | Three quarters | Days | 112 | - | 197 | 197 |
| Fraction of population infected by Covid-19 | General population | Percent | 96 | 73 | 40 | 71 |
|  | Low-risk population | Percent | 96 | 73 | 92 | 96 |

**Fig. 2.**
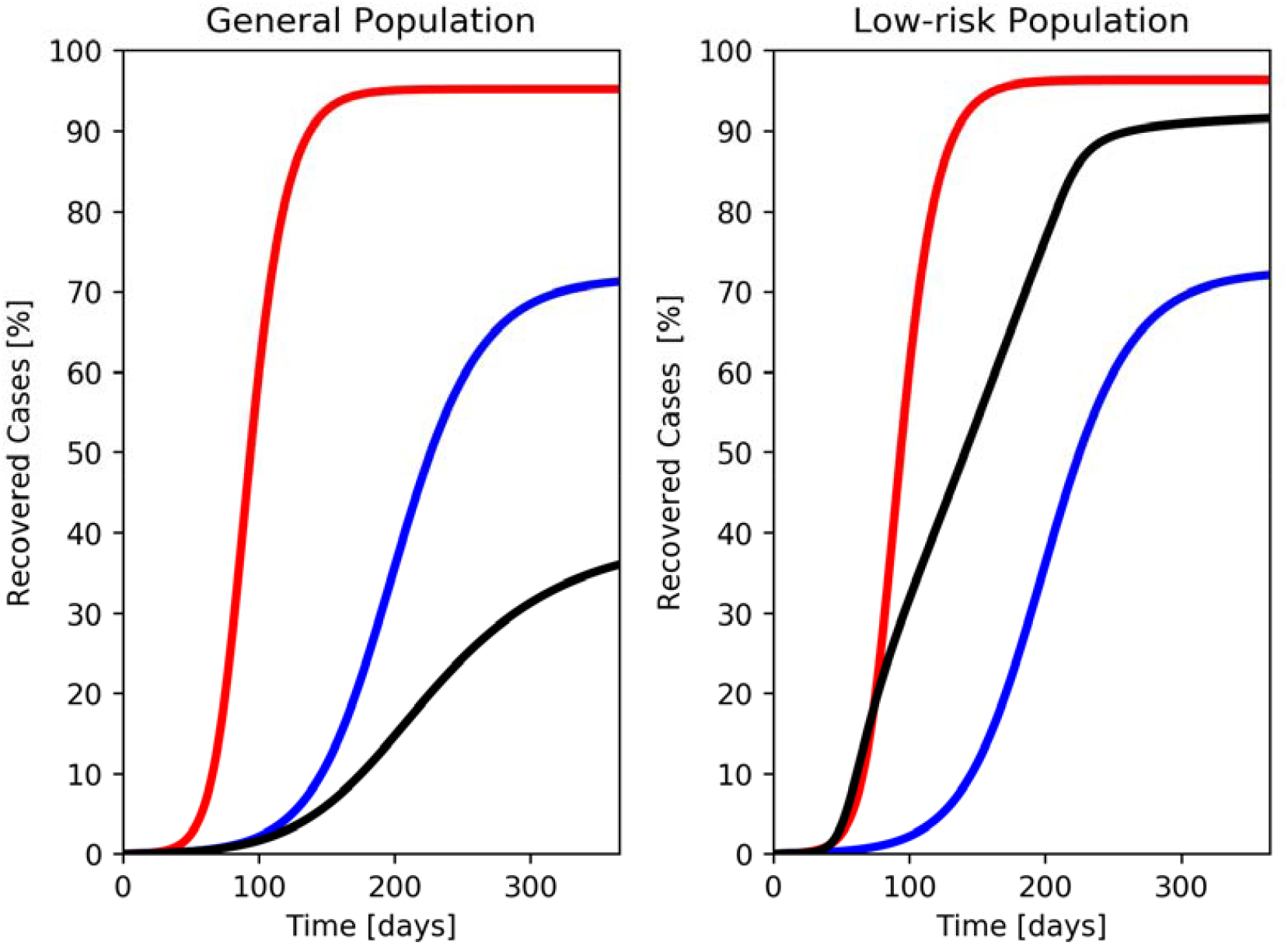
The percentage of recovered cases in three scenarios, in red uncontrolled, in blue mitigation and in black the controlled avalanche strategy. In the left side for the general population and in the right side for the low risk population.

A crucial number in public health management is the number of the intensive care units (ICUs) available. A major challenge is to stay below this limit, which is currently in Israel 2,864 ICUs (25). Fig. 3 shows the ICU hospitalization demand for the four scenarios stated above, where the current limit of ICUs in Israel is marked by horizontal grey dashed line. The two scenarios which include CA implementation exhibit dramatically lower cases in need of ICU hospitalizations. Furthermore, the maxima for the general population in need for ICUs under both CA scenarios is 4,146 cases (and 570 for the low risk population), much lower than the 11,839 and 35,382 cases for mitigation and uncontrolled scenarios, respectively (Table 1).

**Fig. 3.**
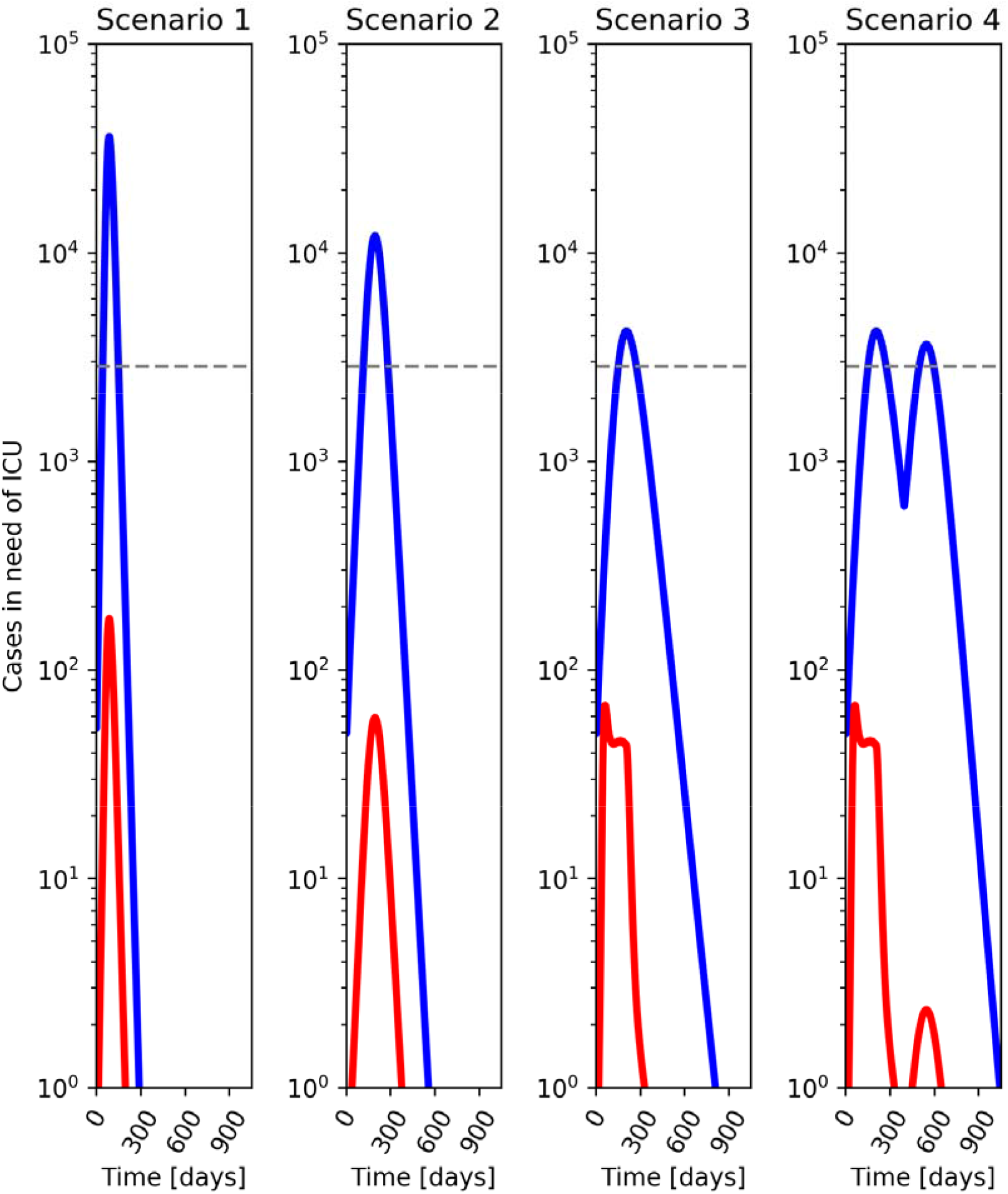
The dynamics of cases in need ICUs for four scenarios: Scenario 1-uncontrolled, Scenario 2 – mitigation, Scenario 3 - controlled avalanche (CA) program, followed by mitigation, Scenario 4 - CA program, followed by uncontrolled policy. The general population is in blue line and those from the low-risk population is in red line.

Although in all scenarios the number of ICU hospitalizations for the total population exceeds the ICU limit in Israel, the magnitude of the surge is dramatically reduced when the CA strategy is implemented. Notably, in all scenarios, the general population is the main source for the ICU demand while the CA volunteers require only very low percentage of the ICU maximal capacity. It should also be pointed out that the analysis is based on the median effective contact rate over 29 provinces, which implemented moderate lockdown in China during early stages of the epidemic (23). More effective lockdown measures as implemented later in China should lead to lower estimates of morbidity and mortality.

## Discussion

In this current reality created by the ongoing Covid-19 epidemic, it is imperative to explore different new control strategies and to rationally evaluate their costs, risks and benefits compared to existing strategies. The proposed CA strategy is not intended to substitute suppression measures, which are extremely important for epidemic containment and prevention of overwhelming available ICU units (26); its efficient implementation is in conjunction with such measures, as early as possible during the initial phase of the epidemic, or as an exit strategy after the initial epidemic has been contained.

Our analysis shows that implementation of the CA strategy is expected to significantly reduce the consequent death toll, to prevent overload of the ICU units through faster formation of overall herd immunity in the population, and to significantly reduce the time period of economic paralysis. The age dependent CFR used in this study does not separate sub-populations with comorbidities. It can be expected that exclusion from the AC program of people that are on the eligible age but suffer from comorbidities, may result in much lower mortality among the volunteers. Furthermore, as the model demonstrates, the main strategy for preventing overload of the ICU, is to increase suppression measures on the general population, and not to limit the CA capacity, whose share of demand for ICU is low. Implementation of the CA policy is thus the safest among all the possible options.

Since implementation depends on voluntary exposure, behavioral, ethical and political considerations should also be considered. As we discuss in detail in the supplementary part, each individual’s decision whether to join the CA program depends on his risk exposure and his individual earning and utility producing opportunities. An individual is more likely to join the program the lower his expected fatality rate from the virus, the higher his probability of contracting the virus if he does not join the program, the higher his benefit from resuming normal life, and the longer he expects the social and economic closure to remain in place.

Our model demonstrates that if 81.5% of people aged 20-_49_ participate in the program, the probability of contracting the virus for non-participants from the general population is 40% if mitigation measures and social distancing are not released after the CA is fully exhausted, and 71% if they are released. It also shows that the CA program is almost fully exhausted after 200 days. If mitigation measures and social distancing are lifted immediately following the end of the program, this implies that anyone not joining the program expects to lose earning opportunities for this period of time. The two variables – alternative risk of exposure to the virus, and time before resumption of normal life – affect the decision whether to join the program. As we explain in the supplementary part, there is some rational expectations equilibrium, such that all people who may join the program, actually choose to do so. The maximum participation age established by such an equilibrium may prove lower than the one analyzed in the model – 50.

Moreover, ethical considerations (also presented in the supplement) may further require lowering the maximum age for participation in the program. Since exposure under the program is voluntary, consent-based criteria support the exposure of young people. Furthermore, since exposed people are isolated, and once they recover and become immune, they rejoin the general population, their exposure reduces negative externalities to other people, and hence public health (and furthermore, social welfare) concerns also support their exposure. The remaining concern is whether the state should impose a limitation on voluntary assumption of risk, due to paternalistic considerations. These considerations do establish a maximum level of risk – or maximum age, above which voluntary exposure cannot be justified. It may therefore well be that the demand for participation in the program would exceed the maximum age allowed. The implementation problem may then be how to prevent higher risk individuals from independently contracting the virus.

To implement CA, governments must have the political will to commit to a program that combines isolation/closure with voluntary exposure for a pre-defined time period. Given extreme suppression measures that governments have implemented, it would seem reasonable that controlled, voluntary exposure could be managed. Rather than opening businesses and public spaces to all low risk groups, thus risking uncontrolled disease spread including to vulnerable populations, governments can leverage success in suppression policies to move to a CA program.

Implementation of the program does not imply that professional personnel will infect people similar to challenge studies. Rather, the government can provide the tools and environment for whoever likes to contract infection to perform it easily and in a regulated manner. Suppression resources may be turned to enforcement of rules regarding screening of applicant volunteers, monitoring mechanisms and patterns of infection, recovery and immunity among volunteers and their social contacts, and regulation of “immunity certification”.

Transparent information regarding CA policy, its uptake, and case studies of volunteers’ experiences should be provided to the public, the media and, in particular, target groups for voluntary exposure. The public, which in many countries has demonstrated its ability to understand and adhere to pandemic related information and edicts, must understand, as much as possible, the risks and benefits of CA to both individuals and society. It would seem they are currently primed to do so (27).

While the anticipated morbidity and mortality among volunteers for CA is expected to be lower than other routinely undertaken risks, government should offer suitable recognition and compensation to volunteers who suffer health setbacks and to their families in case of mortality due to participation. Participants in the program may also be offered to volunteer for studies exploring important questions such as the length of protective immunity. Plasma donated by recovered patients can be used to treat critically ill Covid-19 patients (28, 29). The program may be later adjusted for recruiting volunteers to challenge studies, enhancing the licensure of vaccines, a practice that was recently suggested by several leading scientists and was claimed to be ethically justified in the current situation (30).

Elimination of this particular threat cannot materialize until adequate herd immunity is reached. It is therefore time for policy makers to include herd immunity as the main goal in the array of policy options. The health, economic, and social costs of all the options to achieve this goal need to be more fully evaluated, compared and discussed. These considerations must include the avoidance of getting stuck in one policy direction, sober confronting of the ethical concerns regarding societal attitudes towards risk taking and more complete assessment of the tradeoffs among health impact, economic impact and social capital. This will require a multi sector, professional and representative task force to manage policy options such as CA and the Covid-19 crisis in general.

## Data Availability

Non Relevant

## References

1. Klausner Z, Fattal E, Hirsch E, Shapira SC. A single holiday was the turning point of the COVID-19 policy of Israel. medRxiv. 2020:2020.03.26.20044412.

2. Ferguson N, Laydon D, Nedjati Gilani G, Imai N, Ainslie K, Baguelin M, et al. Report 9: Impact of non-pharmaceutical interventions (NPIs) to reduce COVID19 mortality and healthcare demand. Imperial College London; 2020.

3. Hamzelou J. We have to respect the coronavirus – and learn as the disease evolves 2020 [updated 16/03/2020. Available from: https://www.newscientist.com/article/2237493-we-have-to-respect-the-coronavirus-and-learn-as-the-disease-evolves/#ixzz6IBDr9uAO.

4. Humanity tested. Nature Biomedical Engineering. 2020.

5. Bai Y, Yao L, Wei T, Tian F, Jin DY, Chen L, et al. Presumed Asymptomatic Carrier Transmission of COVID-19. JAMA. 2020.

6. Fraser C, Riley S, Anderson RM, Ferguson NM. Factors that make an infectious disease outbreak controllable. Proc Natl Acad Sci U S A. 2004;101(16):6146–51.

7. Zhuang Z, Zhao S, Lin Q, Cao P, Lou Y, Yang L, et al. Preliminary estimating the reproduction number of the coronavirus disease (COVID-19) outbreak in Republic of Korea and Italy by 5 March 2020. medRxiv. 2020:2020.03.02.20030312.

8. Liu L, Xie J, Sun J, Han Y, Zhang C, Fan H, et al. Longitudinal profiles of immunoglobulin G antibodies against severe acute respiratory syndrome coronavirus components and neutralizing activities in recovered patients. Scand J Infect Dis. 2011;43(6-7):515–21.

9. Wu LP, Wang NC, Chang YH, Tian XY, Na DY, Zhang LY, et al. Duration of antibody responses after severe acute respiratory syndrome. Emerg Infect Dis. 2007;13(10):1562–4.

10. Guo X, Guo Z, Duan C, Chen Z, Wang G, Lu Y, et al. Long-Term Persistence of IgG Antibodies in SARS-CoV Infected Healthcare Workers. medRxiv. 2020.

11. Bao L, Deng W, Gao H, Xiao C J. L, Xue J, et al. Reinfection could not occur in SARS-CoV-2 infected rhesus macaques. medRxiv. 2020.

12. WHO. Report of the WHO-China Joint Mission on Coronavirus Disease 2019 (COVID19). Geneva; 2020.

13. Woelfel R, Corman VM, Guggemos W, Seilmaier M, Zange S, Mueller MA, et al. Clinical presentation and virological assessment of hospitalized cases of coronavirus disease 2019 in a travel-associated transmission cluster. medRxiv. 2020.

14. ECDC. Novel coronavirus (SARS-CoV-2). Discharge criteria for confirmed COVID-19 cases – When is it safe to discharge COVID-19 cases from the hospital or end home isolation?. 2020.

15. Verity R, Okell LC, Dorigatti I, Winskill P, Whittaker C, Imai N, et al. Estimates of the severity of COVID-19 disease. Lancet. 2020:2020.03.09.20033357.

16. Wighton D, Chasan D. Germany will issue coronavirus antibody certificates to allow quarantined to re-enter society 2020 [Available from: https://www.telegraph.co.uk/news/2020/03/29/germany-will-issue-coronavirus-antibody-certificates-allow-quarantined/.

17. Cao Z, Zhang Q, Lu X, Pfeiffer D, Jia Z, Song H, et al. Estimating the effective reproduction number of the 2019-nCoV in China. medRxiv. 2020:2020.01.27.20018952.

18. Read JM, Bridgen JR, Cummings DA, Ho A, Jewell CP. Novel coronavirus 2019-nCoV: early estimation of epidemiological parameters and epidemic predictions. medRxiv. 2020:2020.01.23.20018549.

19. Wu JT, Leung K, Leung GM. Nowcasting and forecasting the potential domestic and international spread of the 2019-nCoV outbreak originating in Wuhan, China: a modelling study. Lancet. 2020;395(10225):689–97.

20. Linton NM, Kobayashi T, Yang Y, Hayashi K, Akhmetzhanov AR, Jung SM, et al. Incubation Period and Other Epidemiological Characteristics of 2019 Novel Coronavirus Infections with Right Truncation: A Statistical Analysis of Publicly Available Case Data. J Clin Med. 2020;9(2).

21. Statistical Abstract of Israel 2019 - No.70. [Internet]. 2019. Available from: https://www.cbs.gov.il/en/publications/Pages/2019/Statistical-Abstract-of-Israel-2019-No-70.aspx.

22. Tourism and Hotel Services Statistics - Quarterly 4 2019. [Internet]. 2019. Available from: https://www.cbs.gov.il/en/publications/Pages/2019/Tourism-and-Hotel-Services-Statistics-Quarterly-4-2019.aspx.

23. Maier BF, Brockmann D. Effective containment explains sub-exponential growth in confirmed cases of recent COVID-19 outbreak in Mainland China. medRxiv. 2020:2020.02.18.20024414.

24. Tian H, Liu Y, Li Y, Wu C-H, Chen B, Kraemer MUG, et al. An investigation of transmission control measures during the first 50 days of the COVID-19 epidemic in China. Science. 2020:eabb6105.

25. Lis J. Israel Has Just 1,437 Ventilators in Reserve for Coronavirus Patients, Lawmakers Told.. Haaretz 2020 26/03/2020.

26. Flaxman S, Mishra S, Gandy A, Unwin H, Coupland H, Mellan T, et al. Report 13: Estimating the number of infections and the impact of nonpharmaceutical interventions on COVID-19 in 11 European countries. Imperial College London (30-03-2020). Imperial College London; 2020.

27. Arundhati Roy: “The Pandemic Is a Portal” 2020 [Available from: https://conversations.e-flux.com/t/arundhati-roy-the-pandemic-is-a-portal/9812.

28. Roback JD, Guarner J. Convalescent Plasma to Treat COVID-19: Possibilities and Challenges. JAMA. 2020.

29. Shen C, Wang Z, Zhao F, Yang Y, Li J, Yuan J, et al. Treatment of 5 Critically Ill Patients With COVID-19 With Convalescent Plasma. JAMA. 2020.

30. Eyal N, Lipsitch M, Smith PG. Human Challenge Studies to Accelerate Coronavirus Vaccine Licensure (Preprint). DASH. 2020.

